# Physical activity and body mass index inequities among adult women in the United States: An application of intersectional multilevel analysis of individual heterogeneity and discriminatory accuracy (I-MAIHDA)

**DOI:** 10.64898/2026.04.06.26350273

**Authors:** Sandra E. Echeverria, Yeongjun Seo, Luisa N. Borrell, Diamond McKelvey, Tala Najjar, Erin J. Reifsteck, Jennifer Toller Erausquin, Jaclyn P. Maher

**Author notes:** **Corresponding Author:** Sandra E Echeverria, PhD, MPH, Dept. Public Health Education, School Health & Human Sciences, The University of North Carolina, Greensboro, 1408 Walker Avenue, Coleman 420-D, Greensboro, NC 27412.

## Abstract

**Background:** Physical activity (PA) and body mass index (BMI) shape cardiovascular risk, particularly in women. Yet, little research exists examining intersectional social axes shaping PA and BMI inequities among women living in the United States (US).

**Methods:** Data included women sampled in the 2015-2020 National Health and Nutrition Examination Survey. We used Intersectional Multilevel Analysis of Individual Heterogeneity and Discriminatory Accuracy (I-MAIHDA) via linear models to examine PA (n=,4591) and BMI (n=4,596) inequities across intersectional strata defined by race/ethnicity, age, education, nativity, and work status. We further quantified the contribution of these strata to the observed inequities and estimated additive fixed effects.

**Results:** In the null model, intersectional strata explained 4.6% and 13.8% of the variance in PA and BMI inequities, respectively, with 99.2% for PA and 97.5% for BMI explained by age, race/ethnicity, education, nativity, and occupation status. On average, Asian and Black women, those aged 35-49 years, those born outside the US, and those with less than a high school diploma had the lowest predicted mean PA. For BMI, Black and Hispanic/Latino women and those younger than 64 years had the highest mean BMI.

**Conclusion:** PA and BMI inequities are mostly explained by race/ethnicity, age, education, nativity, and work status. Our findings offer insights into universal and potential policy-informed health promotion strategies that may be tailored to women with these social identities and lived experiences that have shaped physical activity and body mass index inequities.

**Research Perspective:** *What is new?* To our knowledge, this is the first study to quantitatively investigate the role of intersectional social strata using Intersectional Multilevel Analysis of Individual Heterogeneity and Discriminatory Accuracy (I-MAIHDA) among women in the United States to predict PA behavior, and one of a handful to predict BMI.

*What should be studied next?* Future research should consider additional social axes (e.g., poverty, motherhood) and broader structural conditions (e.g., neighborhood characteristics, policies) that can further elucidate the unique lived experiences of women in the US and how these social axes influence cardiovascular risk factors.

## INTRODUCTION

Physical activity (PA) and body mass index (BMI) are two significant contributors to cardiovascular risk in women. In the United States (US) and around the globe, women of all age groups are less likely to meet recommended levels of PA^1,2^ and they have higher levels of severe obesity^2^ than men. National data in the US indicates that less than 6% of all women of child-bearing age meet proper control of traditional cardiovascular (CV) risk factors.^3^ Given that nearly one in two women will experience cardiovascular disease over their lifetime, it is critical to identify and understand factors that pattern PA and BMI.^4–6^

A large body of evidence has examined how determinants of health, such as race/ethnicity, age, education, nativity, and work conditions, shape PA and BMI. For example, while obesity tends to increase with age for all women, Black women have the highest prevalence of obesity and low levels of PA of all racial and ethnic groups.^6^ Higher educational attainment is associated with increased levels of PA and lower BMI.^7,8^ Moreover, several studies have shown that US-born men and women have a higher prevalence of obesity than their foreign-born counterparts, while leisure-time PA generally increases with longer duration in the US among immigrants.^9–11^ Given that employed adults spend most of their day at work, some evidence indicates that working and type of work have contributed to rising obesity prevalence and lower levels of PA in US men and women.^12,13^ Nevertheless, despite the burgeoning evidence on PA and BMI inequities in women, most of this research has examined social dimensions in isolation, thereby limiting our understanding of the multiple, overlapping social identities that shape women’s lived experiences.

Intersectionality as a theoretical framework offers a critical lens for examining how interlocking systems of power such as sexism, racism, xenophobia, socioeconomic inequality, and ageism structure social inequities in well-being and lived experience.^14^ Intersectionality has its origins in Black feminist thought and community activists who articulated the unique experiences Black women and women of color experience.^15,16^ Moreover, within the past decade and a half, intersectionality has emerged as a key perspective for addressing public health inequities.^17^ In physical activity research, intersectional studies have largely relied on qualitative methods.^18^ Such work offers critical insights that could be complemented by a robust intersectional analysis of large-scale population data. Statistical approaches, such as the intersectional multilevel analysis of individual heterogeneity and discriminatory accuracy (I-MAIHDA), have emerged over the last decade allowing the quantitative examination of the intersection of multiple social identities shaping health inequities.^19^ I-MAIHDA employs multilevel models with an intersectional perspective to examine between- and within-strata variations in health, capturing the clustering of individuals within intersectional social identity strata.^20^ For example, recent work has shown that additive fixed effects of age, sex, educational attainment, nativity, and geographic region used to define intersectional strata explained most of the variance in BMI,^21^ while another study from Spain^22^ using strata of age, sex/gender, immigration status, and education found that older adults, immigrants, and those with lower education experienced the highest BMI means.

These studies highlight additional insights into cardiovascular disease risk that can be gained by employing a quantitative intersectional approach that centers on systems of power and oppression. However, to our knowledge, there has yet to be a study using quantitative approaches like I-MAIHDA to examine intersectional inequities in PA and BMI in a racially and ethnically diverse sample of adult women in the United States. Examining intersectional patterns within two key modifiable risk factors for cardiovascular disease is essential for developing tailored strategies that effectively promote PA and healthy body weight across diverse social axes. Therefore, in the present study, we examine intersectional PA and BMI inequities across race/ethnicity, age, education, nativity, and work status in US adult women, quantify the between-stratum inequities via the variance partition coefficient (VPC), and determine the proportion of variation explained by these social factors among US women. We hypothesize that these social identities are differentially related to high BMI or physically inactivity.

## METHODS

### Study Design and Population

We used data of women aged 20-64 years sampled in the 2015–2016 and 2017–2020 National Health and Nutrition Examination Survey (NHANES). NHANES is a national, probabilistic survey of the non-institutionalized US population that uses a mix of self-reported, examination, and laboratory procedures to collect data. Due to the COVID-19 pandemic, NHANES paused all data collection in March 2020, resulting in a 1.5-year gap in data collection. To address this gap, NHANES provides a merged file combining data collected from the 2017-2018 survey years with data collected between 2018 and March 2020 (labeled ‘pre-pandemic data’). We chose not to merge data from the 2021-2023 cohort and onward with these earlier years given the documented changes in indicators of cardiovascular risk factors, including physical activity and weight gain as a result of the COVID-19 pandemic.^23,24^ Survey measures, datasets, and related documentation are available elsewhere.^25^

A total of 6,736 women aged 20 years or older participated in NHANES during 2015-2016 and 2017-2020. Because of our interest in the role of current employment in shaping participation in modifiable health behavior risk factors such as PA and BMI, we restricted our sample to women 20-64 years of age to capture the largest segment of the working-age population (n=4,793). We excluded women if they were pregnant (n=133) or had missing data on at least one key study variable race/ethnicity, age, education, nativity, or working outside the home (i.e., PA [n=69], BMI [n=364], resulting in an analytic sample size of n=4,591 for PA and n= 4,296 for BMI. Institutional review board approval was not required because the data were deidentified and publicly available.

### Outcomes

Our analysis focused on two outcome variables: PA and BMI. *PA* was assessed using the Global Physical Activity Questionnaire.^26^ Participants were asked to reflect on a typical week and report the number of days and time spent engaging in moderate-to-vigorous intensity PA (MVPA) across different domains (i.e., work, transport, and leisure) on one of the days. Following standard handling of extreme values, participants who reported >960 minutes in a given domain on a typical day were not considered valid responses and were excluded from the analysis. Valid responses were used to create a continuous variable representing the total minutes of MVPA per week. NHANES provides calculated measures of *BMI* based on participants’ weight in kilograms divided by height in meters squared (kg/m^2^). BMI was treated as a continuous variable during the analysis. For both PA and BMI, we report the percent of the population that met 2018 PA guidelines of engaging in 150+ MVPA per week, and those who are obese, overweight, and normal weight status according to CDC classifications when describing our sample population.^27^

### Exposures/Strata Composition

We created intersectional strata based on race/ethnicity, age, education, nativity, and current employment status (work) to capture social identity axes relevant to women’s cardiovascular health. Participants were asked to select a racial category they identify with and if they are of ‘Hispanic, Latino, or Spanish origin’ (herein referred to as Latino). Using NHANES’ classification, women were classified as non–Latina Asian (herein referred to as Asian), non-Latina Black (herein referred to as Black), Latina, and non-Latina White (herein referred to as White). Age was categorized as 20-34, 35-49, and 50-64 years of age. Educational attainment was self-reported by participants (<9^th^ grade, 9-11^th^, high school graduate/equivalent, some college, and college or more) and classified in our study into 2 groups: less than a high school diploma vs high school diploma or higher. We chose these category cutoffs to account for differences in educational attainment by country of origin. Participants were classified as US-born or not based on their reported country of birth. Finally, employment status was determined based on whether participants reported working in the past week, regardless of employment type. The intersection of these social factors resulted in 86 strata due to some missing data in select strata. Each stratum was assigned a unique identifier, with each digit of the strata representing categories of the variables included: Digit 1= race/ethnicity (1=Asian, 2=Black, 3=Latina, 4=White); Digit 2 = age (1=20-34, 2=35-49, 3=50-64); Digit 3 = educational attainment (1=less than high school, 2=high school diploma or above); Digit 4 = nativity (1=US-born, 2 = born outside the US); and Digit 5 = employment status (1=worked in the past week, 2=did not work in the past week).

### Statistical Analysis

Descriptive characteristics were generated for each component of the social strata and presented separately for each outcome. Means and standard deviations (SD) were calculated for continuous variables, and frequencies and percentages were calculated for categorical variables. The number and percentage of participants in each intersectional stratum were determined. For both sets of models, strata with fewer than 10 cases were retained, as I-MAIHDA approaches are designed to accommodate sparsely populated strata through partial pooling, where estimates are adjusted and penalized toward the grand mean to reduce instability and bias associated with sample size. ^20^

Following the methodology developed by Evans et al. (2024),^20^ we used I-MAIHDA via a multilevel linear model, where individuals at level 1 were clustered within intersectional strata at level 2. We first fit an empty (null) random intercept model (Model A) to estimate the between-stratum variance in PA or BMI, thereby capturing stratum-level inequities. To estimate the amount of variation between strata, we calculated the Variance Partition Coefficient (VPC) as follows:

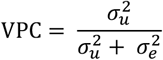

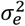represents between individual variance, where 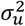 represents between-stratum PA/BMI differences (i.e.,inequities). This measure is calculated as the proportion of the total variance corresponding to level 2, or the between-stratum variance out of the total variance. The second model (Model B) added the variables represented by our intersectional strata as level 2 additive main fixed effects to estimate the average effects of these variables across strata and generate their predicted mean values. The predicted means were used to examine inequities in PA or BMI associated with race/ethnicity, age, educational attainment, nativity, and work status.

To determine the extent to which inequities were explained by the additive fixed effects, we calculated the Proportional Change in Variance (PCV) as follows:

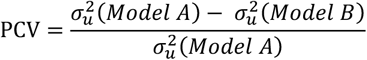

The unexplained variance was considered a deviation from the additive fixed effects or due to intersectional interaction effects.

Data management and statistical analyses were conducted using *SAS v9.4* drawing on the SAS syntax associated with the I-MAIHDA tutorial developed by Borrell (2025).^28^ Similar to previous research,^22,29^ survey weights were not considered in the analyses, and therefore, no generalization inference was made to the U.S. population

## RESULTS

### Study Population

The sample characteristics are shown in **Table 1**. Generally, the characteristics of the sample were consistent across the samples used for PA and BMI models. Therefore, we present descriptions of the sample used for the PA analyses. The largest racial and ethnic group in the sample identified as Latina (29.1%), and the smallest group identified as Asian (15.4%). Participants were distributed across three age groups, with the largest proportion aged 50-64 years (38.18%). Most of the sample had education beyond high school (82.2%), were born in the U.S. (64.4%), and reported working in the past week (60.9%). Approximately one-third (32.0%) and half (51.8%) of the sample met or exceeded the 2018 Physical Activity Guidelines of engaging in 150+ MVPA. More than one-quarter of the sample was classified as underweight or normal weight (28.8%) or overweight (26.5%). Approximately half of the sample (44.7%) was classified as obese.

**Table 1.**
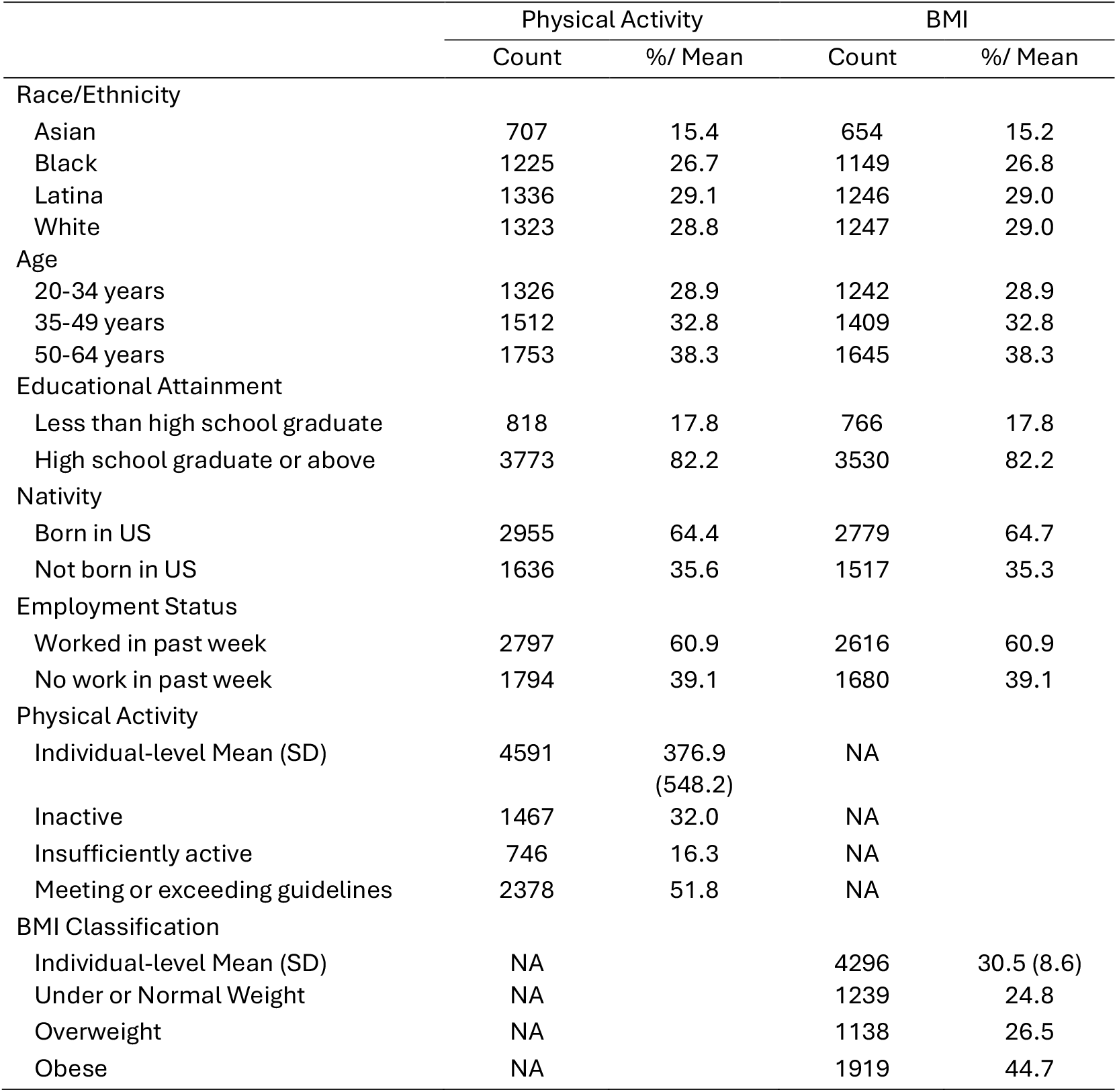
Descriptive Statistics for Women Ages 20-64 by PA and BMI, NHANES 2015-2020.

|  | Physical Activity |  | BMI |  |
| --- | --- | --- | --- | --- |
|  | Count | %/ Mean | Count | %/ Mean |
| **Race/Ethnicity** |  |  |  |  |
| Asian | 707 | 15.4 | 654 | 15.2 |
| Black | 1225 | 26.7 | 1149 | 26.8 |
| Latina | 1336 | 29.1 | 1246 | 29.0 |
| White | 1323 | 28.8 | 1247 | 29.0 |
| **Age** |  |  |  |  |
| 20-34 years | 1326 | 28.9 | 1242 | 28.9 |
| 35-49 years | 1512 | 32.8 | 1409 | 32.8 |
| 50-64 years | 1753 | 38.3 | 1645 | 38.3 |
| **Educational Attainment** |  |  |  |  |
| Less than high school graduate | 818 | 17.8 | 766 | 17.8 |
| High school graduate or above | 3773 | 82.2 | 3530 | 82.2 |
| **Nativity** |  |  |  |  |
| Born in US | 2955 | 64.4 | 2779 | 64.7 |
| Not born in US | 1636 | 35.6 | 1517 | 35.3 |
| **Employment Status** |  |  |  |  |
| Worked in past week | 2797 | 60.9 | 2616 | 60.9 |
| No work in past week | 1794 | 39.1 | 1680 | 39.1 |
| **Physical Activity** |  |  |  |  |
| Individual-level Mean (SD) | 4591 | 376.9<br>(548.2) | NA |  |
| Inactive | 1467 | 32.0 | NA |  |
| Insufficiently active | 746 | 16.3 | NA |  |
| Meeting or exceeding guidelines | 2378 | 51.8 | NA |  |
| **BMI Classification** |  |  |  |  |
| Individual-level Mean (SD) | NA |  | 4296 | 30.5 (8.6) |
| Under or Normal Weight | NA |  | 1239 | 24.8 |
| Overweight | NA |  | 1138 | 26.5 |
| Obese | NA |  | 1919 | 44.7 |

### Exposures/Strata Descriptives

Of the 86 intersectional strata, 63 (73.3%) contained at least 10 cases, and 37.2% had at least 50 cases when predicting PA (**Table 2**). Similarly, for BMI, 59 strata (68.6%) contained at least 10 cases, and 31 strata (36.1%) had at least 50 cases. There was an average of 53 records per stratum (range= 1 - 264) in the PA model and an average of 50 records per stratum (range = 1 - 250) for BMI.

**Table 2.**
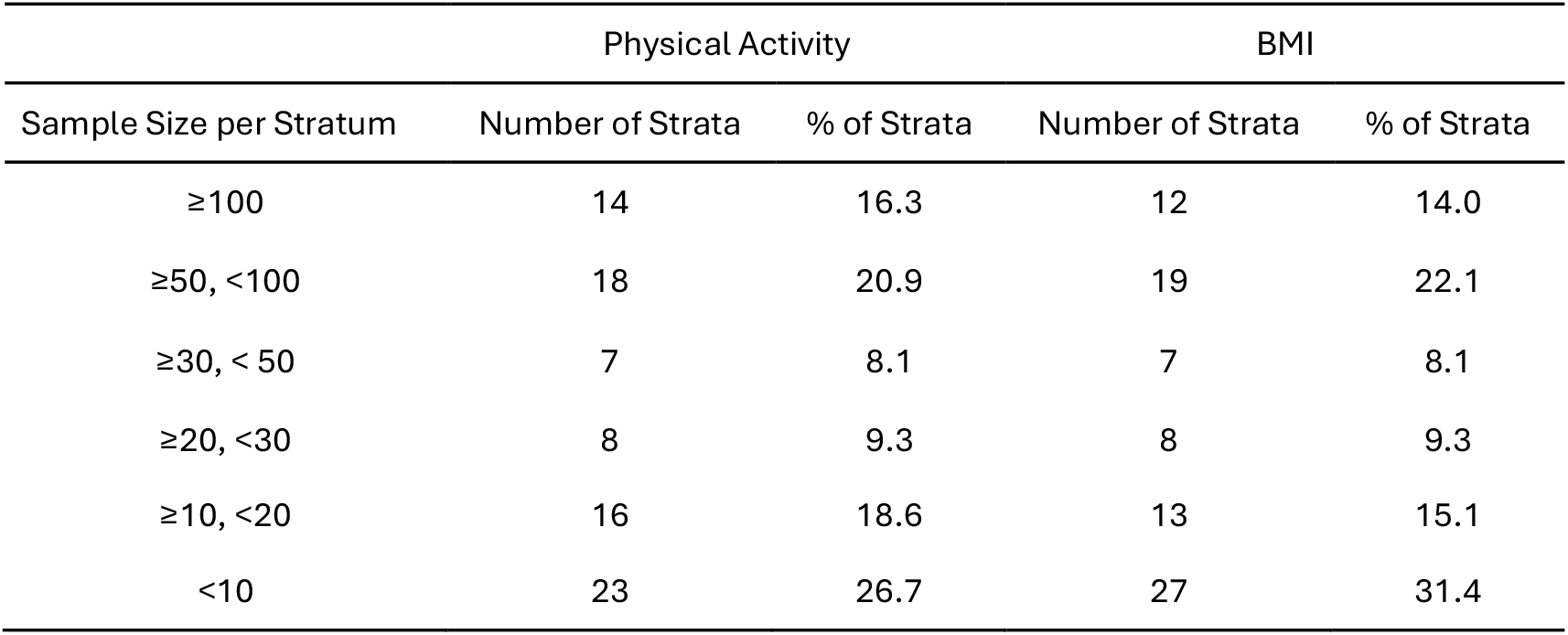
Sample Size of Intersectional Strata for PA and BMI, NHANES 2015-2020.

| Sample Size per Stratum | Physical Activity |  | BMI |  |
| --- | --- | --- | --- | --- |
|  | Number of Strata | % of Strata | Number of Strata | % of Strata |
| ≥100 | 14 | 16.3 | 12 | 14.0 |
| ≥50, <100 | 18 | 20.9 | 19 | 22.1 |
| ≥30, < 50 | 7 | 8.1 | 7 | 8.1 |
| ≥20, <30 | 8 | 9.3 | 8 | 9.3 |
| ≥10, <20 | 16 | 18.6 | 13 | 15.1 |
| <10 | 23 | 26.7 | 27 | 31.4 |

**Table 3** presents the results for the null (Model A) and fixed effects (Model B) I-MAIHDA models for PA and BMI. The null model for BMI had a higher VPC of 13.8%. After including race/ethnicity, age, educational attainment, nativity, and work status as additive fixed effects, the PCV indicated that the additive fixed effects explained nearly all the variation in PA (99.2%), indicating no meaningful interaction across the intersectional strata and that nearly all strata may follow the general inequity patterns predicted by the additive fixed effects.

**Table 3.** Estimates and their 95% confidence intervals from linear mixed models of body mass index (BMI) and physical activity for women, aged 20 or older, who participated in NHANES (2015-2020)

|  | Physical Activity |  |  |  | BMI |  |  |  |
| --- | --- | --- | --- | --- | --- | --- | --- | --- |
|  | Null Model (Model A) |  | Full Model (Model B) |  | Null Model (Model A) |  | Full Model (Model B) |  |
|  | *b* | 95% CI | *b* | 95% CI | *b* | 95% CI | *b* | 95% CI |
| Fixed Effects |  |  |  |  |  |  |  |  |
| Intercept | 361.0 | (325.9, 396.0) | 489.1 | (428.2, 550.0) | 30.0 | (29.2, 30.8) | 29.5 | (28.4, 30.6) |
| Race (Ref: White) |  |  |  |  |  |  |  |  |
| Asian |  |  | -93.2 | (-157.5, -28.9) |  |  | -3.6 | (-4.7, -2.4) |
| Black |  |  | -63.3 | (-107.2, -19.5) |  |  | 2.9 | (2.1, 3.7) |
| Latina |  |  | 17.6 | (-32.2, 67.4) |  |  | 1.6 | (0.7, 2.5) |
| Age (Ref: 20-34) |  |  |  |  |  |  |  |  |
| 35-49 |  |  | -140.5 | (-182.0, -99.1) |  |  | 2.3 | (1.6, 3.1) |
| 50-64 |  |  | -211.2 | (-251.5, -170.9) |  |  | 2.4 | (1.7, 3.1) |
| Education (Ref: < High School) |  |  |  |  |  |  |  |  |
| ≥ High school graduate |  |  | 88.7 | (43.7, 133.7) |  |  | -0.90 | (-1.6, -0.1) |
| Nativity (Ref: US-Born) |  |  |  |  |  |  |  |  |
| Born outside of US |  |  | -83.7 | (-130.4, -37.0) |  |  | -2.4 | (-3.2, -1.6) |
| Working Status (Ref: Worked in last week) |  |  |  |  |  |  |  |  |
| No work in last week |  |  | -5.3 | (-39.0, 28.5) |  |  | 0.4 | (-1.6, 1.0) |
| Random Effects |  |  |  |  |  |  |  |  |
| Strata-level variance | 14552.0 | (9351, 25730) | 114.4 | (11.3, 7.2) | 10.5 | (7.3, 16.4) | 0.26 | (0.1, 24.0) |
| Individual level variance | 286806 | (275341, 299006) | 286975.0 | (275525, 299157) | 65.4 | (62.7, 68.2) | 65.6 | (62.8, 68.4) |
| Summary Statistics (%) |  |  |  |  |  |  |  |  |
| Variance Partition Coefficient | 4.8 |  | 0.04 |  | 13.8 |  | 0.40 |  |
| Proportional Change in Variance |  |  | 99.2 |  |  |  | 97.5 |  |

Note: The null model is empty, and the full model includes the main additive effect model.

Estimates from Model B indicated significant differences in PA by race/ethnicity, age, education, and nativity status, but not by working status (Table 3). Compared with White women, Asian (*b* = -93.2, 95% Confidence Interval (CI):-157.5,-28.9) and Black (*b* = -63.3, *95% CI: -107.2,-19.5*) women had significantly lower PA estimates. Women aged 35-49 (*b* = -140.53, *95% CI:-182.0,-99.1)* and 50-64 (*b* = -211.2, 95% CI: -251.5,-170.9) engaged in significantly less PA than women aged 20-34. Finally, women with less than a HS diploma, women with a HS diploma or more (*b* = 88.7, *95% CI: 43.7,133.7*) engaged in more PA than those with more than a high school education, whereas women born outside the US (*b* =-83.7, *95% CI: -130.4,-37.0*) engaged in less PA than their US-born counterparts. For models with BMI as the outcome, the PCV indicated slightly higher effects due to interactions (2.5%).

Similar trends were observed regarding model estimates for BMI among racial/ethnic groups, age, education, and nativity status Table 3). When compared with White women, Asian women had a significantly lower BMI (*b* = -3.6, 95% CI: -4.7,-2.4), whereas Black (*b* = 2.9, 95% CI: 2.1,3.7) and Latina (*b* = 1.6, *95% CI: 0.7,2.5*) women had significantly higher BMI values. Women aged 35-49 (*b* = 2.3, 95% CI: 1.6,3.1) and 50-64 (*b* = 2.4, 95% CI: 1.7,3.1) had significantly higher BMI values than men aged 20-34. Women with a HS diploma or more (*b* = -0.9, *95% CI: -1.6,-0.1*) had significantly lower BMI values than those with less than a HS diploma. Women born outside the US (*b* = -2.4, 95% CI: -3.2,-1.6) had significantly lower BMI than those born in the US. There were no differences among women’s BMI by working status (*p* = 0.15).

### Intersectional social strata

The predicted PA means for the strata ranked from low to high are displayed in **Figure 1a**. A total of 53.7% of PA means fell below the intercept mean of 360.98 (Model A). **Table 4** presents the strata with the highest (514 - 596 minutes) and lowest (96 to 166 minutes) predicted PA per week. Consistent with the additive fixed effects, the highest predicted levels of PA were observed in Latina women aged 20-34 years with a high school diploma, born in the U.S., and working, and the lowest predicted values were observed among Asian women aged 50-64 years without a high school education, born in the US, and not working. The difference between the highest and lowest predicted PA means was 500 mins, which is substantially higher than recommended guidelines of meeting 150+ of MVPA/week.

**Table 4.** Highest and Lowest ranked strata for predicted mean physical activity and body mass index among women 20-64 years of age, NHANES 2015-2020.

| Highest Physical Activity Strata |  |  |  |  |  |  | Lowest Physical Activity Strata |  |  |  |  |  |  |
| --- | --- | --- | --- | --- | --- | --- | --- | --- | --- | --- | --- | --- | --- |
| N | Race/<br>Ethnicity | Age | ≥ HS<br>Graduate | Born in<br>US | Work-<br>ing | Predicted Mean<br>(95 % CI) | N | Race/<br>Ethnicity | Age | ≥ HS<br>Graduate | Born in<br>US | Work-<br>ing | Predicted Mean<br>(95 % CI) |
| 136 | Latina | 20-34 | Yes | Yes | Yes | 595.8 (546.4, 645.2) | 24 | Asian | 50-64 | No | No | No | 95.8 (33.6, 158.1) |
| 53 | Latina | 20-34 | Yes | Yes | No | 589.2 (535.1, 643.2) | 19 | Asian | 50-64 | No | No | Yes | 101.3 (37.0, 165.7) |
| 264 | White | 20-34 | Yes | Yes | Yes | 576.6 (535.1, 618.1) | 6 | Black | 50-64 | No | No | No | 125.9 (60.7, 191.1) |
| 104 | White | 20-34 | Yes | Yes | No | 576.4 (528.9, 623.9) | 3 | Black | 50-64 | No | No | Yes | 130.9 (63.6, 198.2) |
| 199 | Black | 20-34 | Yes | Yes | Yes | 514.2 (470.9, 557.6) | 14 | Asian | 35-49 | No | No | No | 166.4 (102.6, 230.2) |
| Highest BMI Strata |  |  |  |  |  |  | Lowest BMI Strata |  |  |  |  |  |  |
| N | Race/<br>Ethnicity | Age | ≥ HS<br>Graduate | Born in<br>US | Work-<br>ing | Predicted Mean<br>(95 % CI) | N | Race/<br>Ethnicity | Age | ≥ HS<br>Graduate | Born in<br>US | Work-<br>ing | Predicted Mean<br>(95 % CI) |
| 27 | Black | 20-34 | No | Yes | No | 35.1 (34.0, 36.4) | 73 | Asian | 20-34 | Yes | No | No | 23.1 (22.0, 24.2) |
| 28 | Black | 50-64 | No | Yes | Yes | 34.7 (34.0, 36.2) | 64 | Asian | 20-34 | Yes | No | Yes | 23.4 (22.3, 24.6) |
| 15 | Black | 50-64 | No | Yes | No | 34.8 (33.4, 36.1) | 1 | Asian | 20-34 | No | No | No | 23.5 (22.1, 25.0) |
| 172 | Black | 50-64 | No | Yes | Yes | 35.0 (34.0, 36.0) | 8 | Asian | 20-34 | No | No | Yes | 24.0 (23.0, 25.4) |
| 73 | Black | 20-34 | No | Yes | No | 34.6 (34.0, 36.0) | 127 | Asian | 35-49 | Yes | No | No | 24.9 (24.0, 26.0) |

Note: Race/ethnicity: Asian, Black, Latina, White; Age: 20-34, 35-49, 50-64; Educational Attainment: Yes, High School Diploma, No, Less than High School Diploma; Born in US: Yes, US Born, No, Born outside the US; Working status: Yes, Worked in the last week, No, Did not work in the last week.

**Figure 1.**
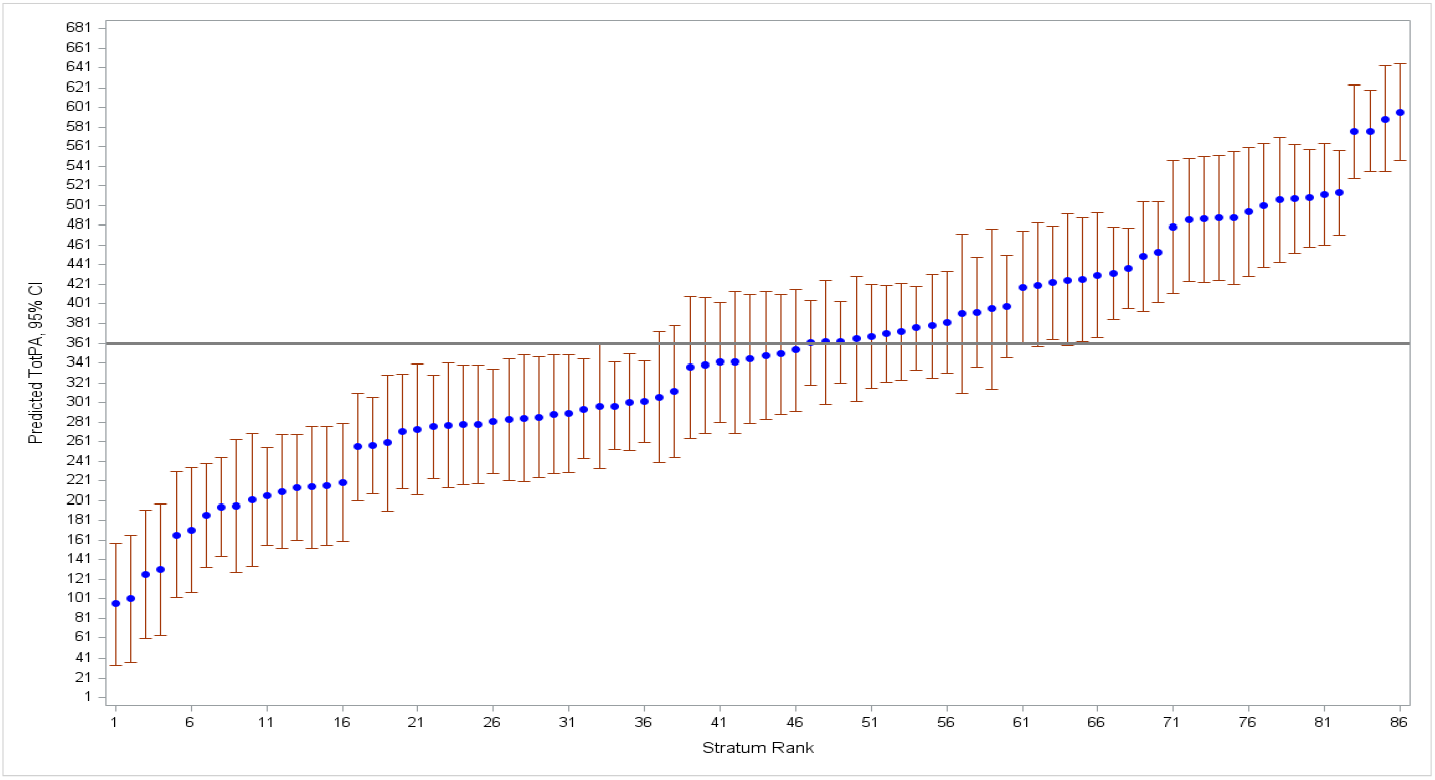
1a.Predicted mean of Total PA rank (Model A) by strata among women 20-64 years of age, NHANES 2015-2020.

The predicted BMI means for the strata ranked from low to high are displayed in **Figure 1b**. In contrast to PA, less than half (44.8%) of the strata means for BMI fell below the intercept value of 30.0. The highest predicted BMI value was observed in Black women aged 20-34 years with a high school education, born in the US and not working (35.13; Table 4). The lowest predicted BMI values were consistently observed for Asian women aged 20-34 born outside the US, with less than a high school education and working status (23.08-24.00). There was a 12-point difference in BMI means between the highest and lowest BMI means. These BMI results are consistent with the PCV observed, where 2.5% of the variance may be explained by interaction effects (see Supplemental Figure 1).

**Figure 1b.**
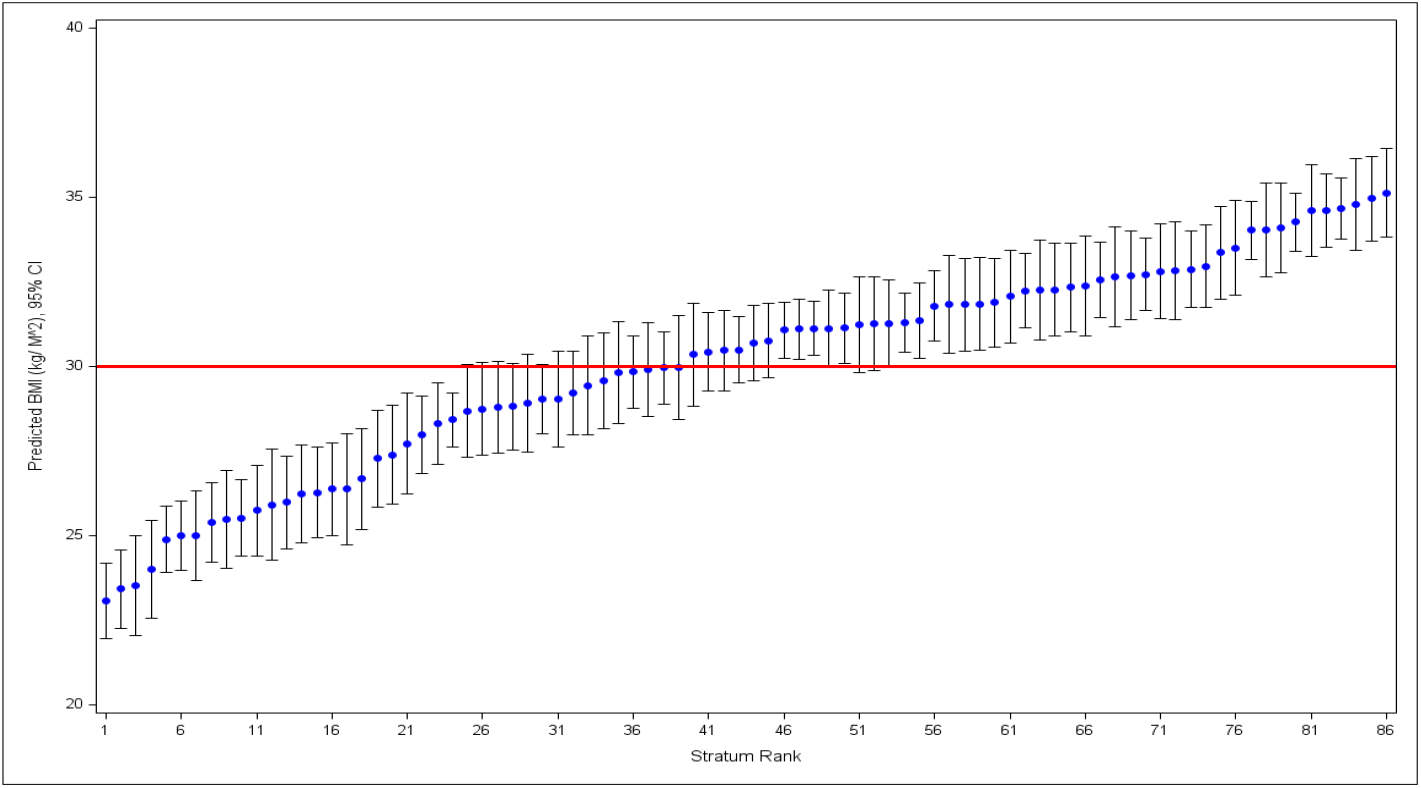
Predicted means of BMI rank (Model A) by strata among women 20-64 years of age, NHANES 2015-2020. NOTE: The predicted means are based on estimates from Model B; the horizontal red line represents the intercepts from Model 1A for each outcome.

## DISCUSSION

We identified intersectional inequities in PA and BMI patterned by social axes of race/ethnicity, age, education, nativity, and work status. Intersectional strata explained a slightly higher proportion of the variance in BMI than in PA, resulting in 2.5% of the variance due to intersectional interactive effects. Although nearly none of the strata deviated from expected patterns for PA, resulting in no intersectionality-defined interaction, we observed some departures from additive fixed effects. Specifically, we observed the highest predicted mean PA for US-born Latina and White women of younger age and higher educational attainment. Similarly, we observed that the 5 lowest estimated mean PA involved Asian and Black women who were born outside the US and had less than a high school diploma. For BMI, the social strata with the 5 highest predicted BMI mean values included Black women with low educational attainment and born in the US, whereas the 5 lowest strata, included Asian women of younger age born outside the US, regardless of educational attainment or work status.

In our study, we leveraged the strengths of I-MAIHDA and found that age, education, nativity and work status contributed to PA in a large racial/ethnic diverse sample of US women. While several studies over the last two decades have examined inequities in cardiovascular risk factors using various modeling techniques (e.g. joint effects, 2 or 3-way interactions, and moderation-mediation models),^30–35^ these studies have not examined intersectional inequities across a large number of socially-defined strata, nor have they focused on women’s cardiovascular risk. Most studies have examined differences in women’s PA by age or education using isolated independent predictors of health.^8,36–39^ Moreover, although results from the additive fixed effect models indicated that our measures of social position explained the majority of the predicted mean in PA and BMI, the intersectional approach leveraged in this study illuminates varied patterns across the 86 strata, pointing to interconnected structural oppressions shaping women’s health risks and behaviors. For example, strata consisting of US-born, young adult, Black women had some of the highest estimates of PA (data not shown). While cautious interpretation is warranted given the small sample size within some individual strata (i.e., approximately 30% comprised of less than 10 women), these results highlight the value of an intersectional approach for capturing heterogeneity in PA patterns across social strata, extending beyond previous research that has typically treated racially and ethnically minoritized women as homogeneous groups (Lee et al., 2023).^40^

Results for physical activity also raise intriguing questions for future intersectional research. We found that Latina and White women were ranked in the top strata of PA. However, this finding may reflect the use of a measure of total PA rather than domain-specific PA. Among US women, differences emerge when considering PA accrued during leisure vs. work (occupational PA). For example, Latina women tend to engage in more occupation-related PA whereas White women tend to engage in more leisure-time PA.^41^ Therefore, examining differential effects of domain-specific PA from a quantitative intersectional approach is an important direction for future research, especially considering emerging evidence that occupational PA is associated with more adverse health outcomes related to cardiovascular risk.^42^

In contrast to PA research, several studies have examined intersectional inequities in BMI using I-MAIHDA. Similar to our findings, Anindya et al (2025) found that the additive fixed effects components included in their intersectional strata (i.e., age, sex, educational attainment, nativity, and geographic region) explained most of the variance in the outcome, resulting in minimal or no interaction effects.^21^ In another study based in Spain, Borrell et al.^22^ found that significant inequities in women’s BMI values emerged when considering intersectional dimensions of age, education, and immigration status. Immigrant women, particularly older women, experienced the highest BMI risk, regardless of education and survey year. Findings from our study indicated the strata that include Asian women born outside of the US with low levels of education tended to have the lowest estimates of BMI, while strata including US-born Black women with lower education had the highest estimated BMI, regardless of age.

The present study has several limitations that warrant attention. First, the cross-sectional nature of this data is unable to provide information on the dynamic nature of social processes and how they influence health over time. Second, although NHANES collects data using validated measures that are strongly linked with cardiovascular disease risk, there are limitations of these measures. Contextual and cultural factors, such as social structures and language, may shape responses to self-reported PA. For example, diversity in nativity and education, translation issues or lack of culturally appropriate items could compromise the validity of reported PA levels.^43^ PA was also assessed using self-reported measures with the potential for measurement error when compared to device-based assessment of PA.^44^ However, self-reported PA remains the most reliable and feasible way to assess active living in large-scale population samples.^45,46^ Relatedly, issues regarding BMI as a measure of health have been debated as BMI values can over- or underestimate body fat and cardiometabolic risk depending on sex and race/ethnicity.^47,48^ This underscores a need for culturally responsive measurement approaches that are able to capture lived realities of marginalized groups in ways that center their experiences in research with an eye toward critical praxis. ^17,49^ Finally, we may have miscategorized the variables included in the strata definition, and that may have affected our findings. The same applies to the systems we intend these variables to represent.

Despite potential limitations, our study has notable strengths and points to potential public health interventions. An important strength of the study is that data were drawn from NHANES to capture health patterns in a large and diverse sample of women in the US. Leveraging I-MAIHDA, a comprehensive approach was used to examine multiple intersecting social axes including race/ethnicity, age, education, nativity and work status to better understand how these factors are associated with cardiovascular health, considering both PA and BMI. We also used clinical assessments of BMI avoiding potential biases with self-reported measures. Our findings also inform potential policy and intervention-related approaches that prioritize health inequities across multiple social dimensions. Specifically, policies and interventions aimed at promoting women’s cardiovascular health should consider the unique needs of women with specific intersecting social axes. Moreover, although PA and BMI were largely explained by additive fixed effects, our findings emphasize the importance of considering multiple cardiovascular risk factors as strata patterns varied for each outcome. Results suggest the need to treat each indicator of cardiovascular risk as a unique outcome and avoid assumptions that socially defined strata apply to all cardiovascular outcomes equally. Investigating strata associated with unique cardiovascular risk factors in women is an important direction for future research. Health advocates and practitioners must balance the tailored nature of health promotion efforts with potential universal applicability to create effective yet scalable interventions targeting at-risk groups.

This study identified intersectional inequities in PA engagement and BMI across social axes defined by race/ethnicity, age, education, nativity, and work status. To our knowledge this is the first study to quantitatively investigate the role of intersectional social strata to predict PA behavior and one of only a handful to predict BMI among women living in the US. Through this approach, we were able to identify some consistent findings in cardiovascular indicators across social strata. There were also unique differences in social strata when predicting PA compared to BMI. Although further work in this line of inquiry is needed, our findings serve as foundational knowledge to understand and intervene on multiple, underlying social determinants of health with respect to the unique lived experiences of women from diverse backgrounds. Thus, our findings can inform potential universal health promotion and intervention strategies tailored to specific subgroups of women at risk.^50^

## Data Availability

We used daat from the National Health and Nutrition Examination Study (NHANES), a publicly avaiable data source sponsored under the National Center for Health Statistis.

https://wwwn.cdc.gov/nchs/nhanes/

---

**Supplemental Figure 1.**
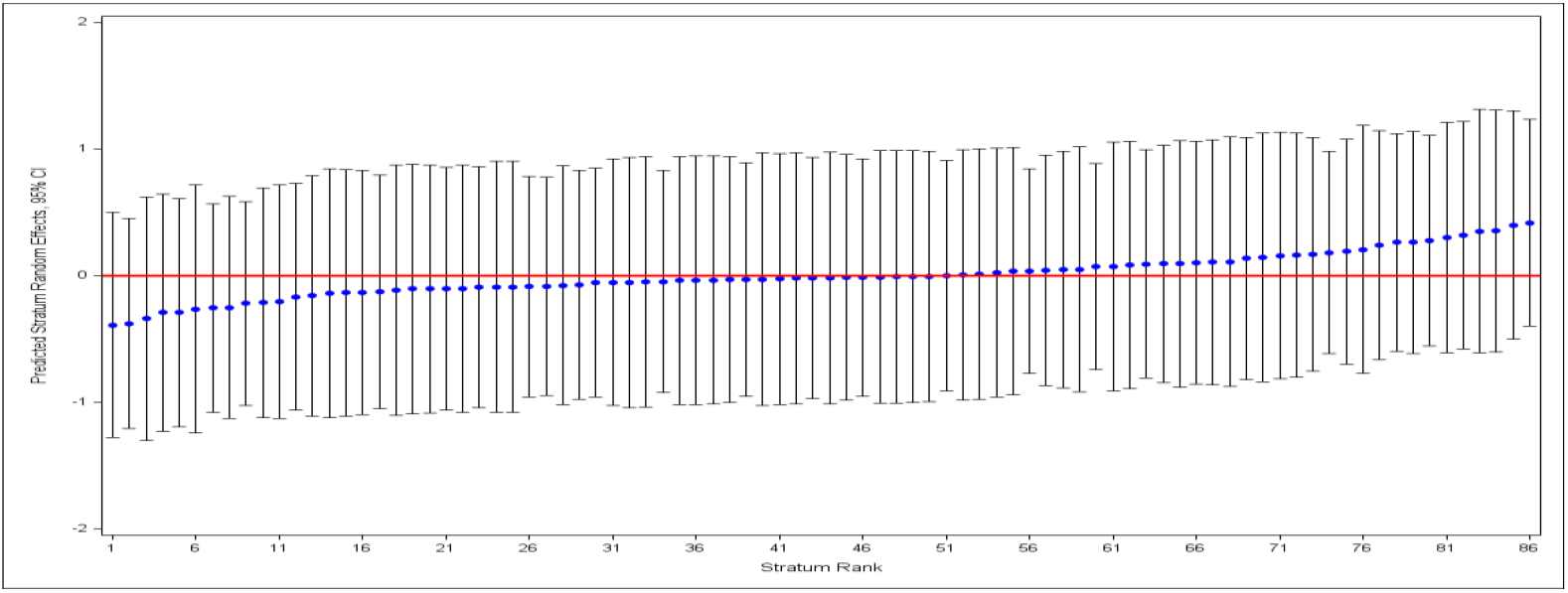
Predicted stratum random effects of BMI among women 20-64 years of age, NHANES 2015-2020.

